# Clinical Phenotypes of Post-Acute Sequelae of SARS-CoV-2 (PASC) Infection: A Longitudinal Cohort Study from Karachi, Pakistan

**DOI:** 10.64898/2026.09.04.26361413

**Authors:** Iffat Khanum, Kumail Ahmed, Tahir Munir, Haniya Zia, Wajiha Saif, Haleema Sadia, Syed Faisal Mahmood, Nosheen Nasir, Kulsoom Ghias, Peter Rabinowitz, Wesley Van Voorhis, Najeeha Talat Iqbal

**Affiliations:** Department of Medicine, Aga Khan University, Karachi, Pakistan; Department of Pediatrics and Child Health, Aga Khan University, Karachi, Pakistan; Department of Biological and Biomedical Sciences; Depts. of Environmental/Occupational Health Sciences, Family Medicine, Global Health, University of Washington, Seattle, Washington, United States of America; Center for Emerging and Re-emerging Infectious Diseases (CERID), University of Washington, Seattle, Washington, United States of America

## Abstract

Post-Acute Sequelae of SARS-CoV-2 infection (PASC), commonly referred to as Long COVID, is increasingly recognized as a significant public health concern. However, longitudinal studies examining its clinical course remain limited in South Asia, and the true burden of persistent symptoms in Pakistan is not well established. This study aimed to characterize the long-term clinical manifestations of PASC and investigate the relationship between symptom persistence, faecal viral shedding, and serotonin levels over a one-year follow-up period.

We conducted a prospective longitudinal study involving individuals diagnosed with PASC (n=60). Participants underwent serial assessments for clinical symptoms, stool SARS-CoV-2 RNA detection, and serotonin measurements in platelets rich plasma at multiple time points over 12 months. A symptom score was developed using 22 symptoms reported by more than 30% of participants.

Among the study population, 60% were female, with a mean age of 42.1 ± 12.1 years. Two-thirds (66%) were overweight or obese, and 28.3% reported multiple SARS-CoV-2 reinfections. Longitudinal analysis demonstrated a significant reduction in overall symptom burden at 9 months (β = −2.68, 95% CI: −3.81 to −1.55; p < 0.001) and 12 months (β = −3.00, 95% CI: −4.13 to −1.87; p < 0.001) compared with baseline. Despite this overall improvement, several symptoms persisted throughout follow-up. The prevalence of myalgia remained largely unchanged (60% at baseline vs. 63% at 12 months), while musculoskeletal symptoms persisted at similar levels (59% vs. 60%). Neurocognitive symptoms declined from 40% to 31.3%, whereas psychological symptoms increased slightly from 40% to 45.8% over the study period.

Faecal viral shedding was detected in 56.7% of participants and demonstrated both intermittent and persistent patterns. Although no association was observed between faecal shedding and symptoms at baseline, significant associations emerged at six months, particularly for breathlessness, fatigue, weakness (77.8%), and joint pain (88.9%). Serotonin levels remained broadly comparable to those reported during acute COVID-19 infection.

These findings highlight the prolonged nature of PASC, with persistent symptoms such as myalgia, joint pain, weakness, and fatigue continuing for at least one year after infection. The persistence of symptoms, together with evidence of ongoing faecal viral shedding, suggests that viral persistence and sustained physiological disturbances may contribute to the long-term pathogenesis of Long COVID in a subset of affected individuals.

## Introduction

The impact of COVID-19 beyond acute infection affected approximately 400 million individuals around the globe, with a health condition known as Post Acute Sequelae of COVID or PASC (1, 2). World Health Organization (WHO) defined PASC as the continuation or development of new symptoms 3 months after the initial SARS-CoV-2 infection, lasting for at least 2 months, with no other alternative explanation (3). This chronic illness can range from mild impairment to a severe, debilitating condition affecting multiple organ systems (4, 5).

The term PASC was introduced by the National Institutes of Health (NIH) as a disease with heterogeneous symptoms and a diverse range of long-term effects that occur following SARS-CoV-2 infection(6). Likewise, the Centers for Disease Control and Prevention (CDC) uses the term Post-COVID Conditions (PCC) to describe a broad spectrum of physical, psychological and social health consequences that continue or emerge four weeks or more after the initial infection (3, 7). Clinically, PCC encompasses a broad spectrum of symptoms affecting multiple organ systems and is therefore characterized as a chronic, multi-system condition(8). To study different domains of PASC, NIH developed a consortium “RECOVER” (Researching COVID to Enhance Recovery), that reported a constellation of 37 most common symptoms including post exertional malaise (PEM), fatigue, brain fog, loss of smell, gastrointestinal symptoms (9, 10), chronic cough, and chest pain (6). Other frequently reported manifestations of PASC in other cohorts include neuropsychiatric symptoms (5) such as depression and anxiety, cough and shortness of breath (11), and cardiovascular symptoms (12).

The proposed pathophysiological mechanism underlying PASC or PCC is interconnectivity of major organ systems affecting the autonomic nervous system, affecting cardiovascular and respiratory systems, which leads to hypoxia-related musculoskeletal disorder (13), and microvascular damage due to chronic inflammation (14). The persistence of viral components in tissues (15) leads to immune dysregulation (16, 17) and autoimmunity (18). Additional factors such as mitochondrial dysfunction (19), oxidative stress, gut dysbiosis (20), and autonomic nervous system disturbances may further contribute to the persistence of symptoms and complications following recovery from acute COVID-19 infection.

PASC has not been systematically studied in LMIC settings where the burden of communicable and non-communicable diseases is disproportionately high, and many patients with Long COVID are living with chronic debilitating conditions in these settings (21–23). To date, no published studies have reported longitudinal data on PASC from Pakistan, highlighting a dire need for longitudinal trajectories of PASC in LMIC (24). Therefore, we report the first longitudinal study examining PASC symptoms from Karachi, Pakistan. Participants were recruited from the hospital and followed over a period of one year to describe the prevalence, persistence, and recovery patterns of PASC symptoms over 12 months.

## Methods

### Study Design and Setting

This was a prospective longitudinal cohort study conducted in collaboration with the United World Antiviral Research Network (UWARN), Pakistan. The study was conducted in a tertiary care setting, Aga Khan University, Karachi, Pakistan. Participants were enrolled through ongoing UWARN-based cohorts using community-based recruitment strategies. Enrolment occurred from March 2023 to July 2024.

Recruited participants were asked to complete a questionnaire to assess retrospective symptoms experienced during acute COVID-19 infection and in the post-COVID period. Participants were then followed prospectively for a period of 12 months. Follow-up assessments were conducted at five time points: baseline, 3 months, 6 months, 9 months, and 12 months.

All follow-up assessments were carried out at Long COVID clinics at the Aga Khan University Hospital, Karachi. At each follow-up visit, participants completed a standardized questionnaire and structured case report forms to document persistent and newly developed symptoms, and blood samples were collected for laboratory analyses. Retention strategies included reminder phone calls and home-based blood sample collection when required. The final follow-up was completed in June 2025, marking the completion of data collection for the study.

### Ethical Consideration

Ethical approval was obtained from the Ethical Review Committee (ERC) of Aga Khan University prior to the initiation of the study with approval number ERC#4794. Informed consent was obtained from all participants or their legal guardians.

### Study Population

#### Inclusion Criteria

The study included adult participants aged >18 years who were permanent residents of Karachi.

All participants included in the study had evidence of prior SARS-CoV-2 infection at some point in their lives, confirmed by a positive COVID-19 PCR and/or antibody test, and were experiencing post-COVID conditions (PCC) as described by the Centre for Disease Control and Prevention (CDC), defined as persistent signs and symptoms lasting more than 4 weeks after the acute phase of infection (19).

#### Outcome Measures and Statistical Analysis

For statistical data analyses, R (version 4.3.2; R Foundation for Statistical Computing, Vienna, Austria) was used. Normality for quantitative variables, such as age, BMI, vital parameters, and other variables, was first evaluated with the Shapiro-Wilk test. For normally distributed variables, mean (SD) was shown, whereas non-normally distributed variables were expressed as median (IQR). For categorical variables, such as gender, number of COVID-19 infections, BMI categories, comorbidities, COVID-19 history and vaccine doses, tobacco use and smoking status, frequencies or proportions were used.

The structured questionnaire included symptoms covering the most reported PASC manifestations. Symptoms were categorized into constitutional (fatigue, weakness, weight changes, recurrent fever), neurocognitive (brain fog/fuzzy brain, memory difficulties, concentration problems, decision-making difficulties), psychological (anxiety, low mood, flashbacks), sleep-related (sleep disturbances, nightmares), headache (frequent headaches, severe headaches, base of skull pain), neuro-ophthalmologic (visual disturbances), respiratory (cough, breathlessness, nasal issues, voice issues, throat issues, throat restriction), cardiovascular (chest pain, palpitations), olfactory/gustatory (anosmia, ageusia, metallic taste), otolaryngological (tinnitus), musculoskeletal (myalgia, joint pain), dermatological (severe rash, frequent rash), and gastrointestinal (nausea) domains (Suppl.Table S1).

For composite score calculation at each time point, positive symptoms were added as a cumulative score. Each symptom was assigned a value of “1” if the symptom was present or “0”, if the symptom was absent. The total cumulative score showed a composite score for each patient. The total range of symptoms was calculated in the range of 0 to 22, where higher scores indicated greater symptom positivity. Symptom positivity analyses were expressed as proportions and visualized using radar plots for distribution at each time point. Hierarchical Clustering was also performed for similar symptoms grouped together in longitudinal data points. The significance of the symptom scores over time was analysed using the Wilcoxon Signed Rank Test.

For longitudinal data analysis, a Linear Mixed-Effects Regression Model was applied to assess random effects where participants were sampled multiple times, and changes at the individual level to account for any within-person correlation. Time was modelled as a categorical variable, with the baseline serving as the reference group, and a general time effect comparing baseline at different time points. The covariates used in the model were age, sex, categories of BMI, the number of times infected with COVID-19, hospitalization, presence of comorbidity, vaccination status, days from infection, and smoking status. The effect sizes are reported as β-coefficients with a 95% CI. All tests were two-tailed, and statistical significance was set at p<0.05.

For biomarker (Serotonin) analyses, the median values were compared using the Kruskal-Wallis test . All statistical analyses were conducted using R (version 4.3.2; R Foundation for Statistical Computing, Vienna, Austria)

### .Biological Sample collection

Blood and stool samples were collected from enrolled participants at baseline and during follow-up visits (6 and 12 months). Blood samples were drawn into EDTA tubes, while stool samples were collected using DNA/RNA Shield Fecal Collection Tubes (Cat # R1101, Zymo Research, USA). Upon arrival at the laboratory, the samples were processed in a biosafety cabinet. Details of the tests that were run on biological samples are shown in Figure 1B. Stool samples were aliquoted into four separate cryovials. Plasma was separated from blood samples by centrifugation at 2000 rpm for 10 minutes, resulting in two aliquots. All aliquoted samples were stored at -80°C until further use.

**Figure 1.**
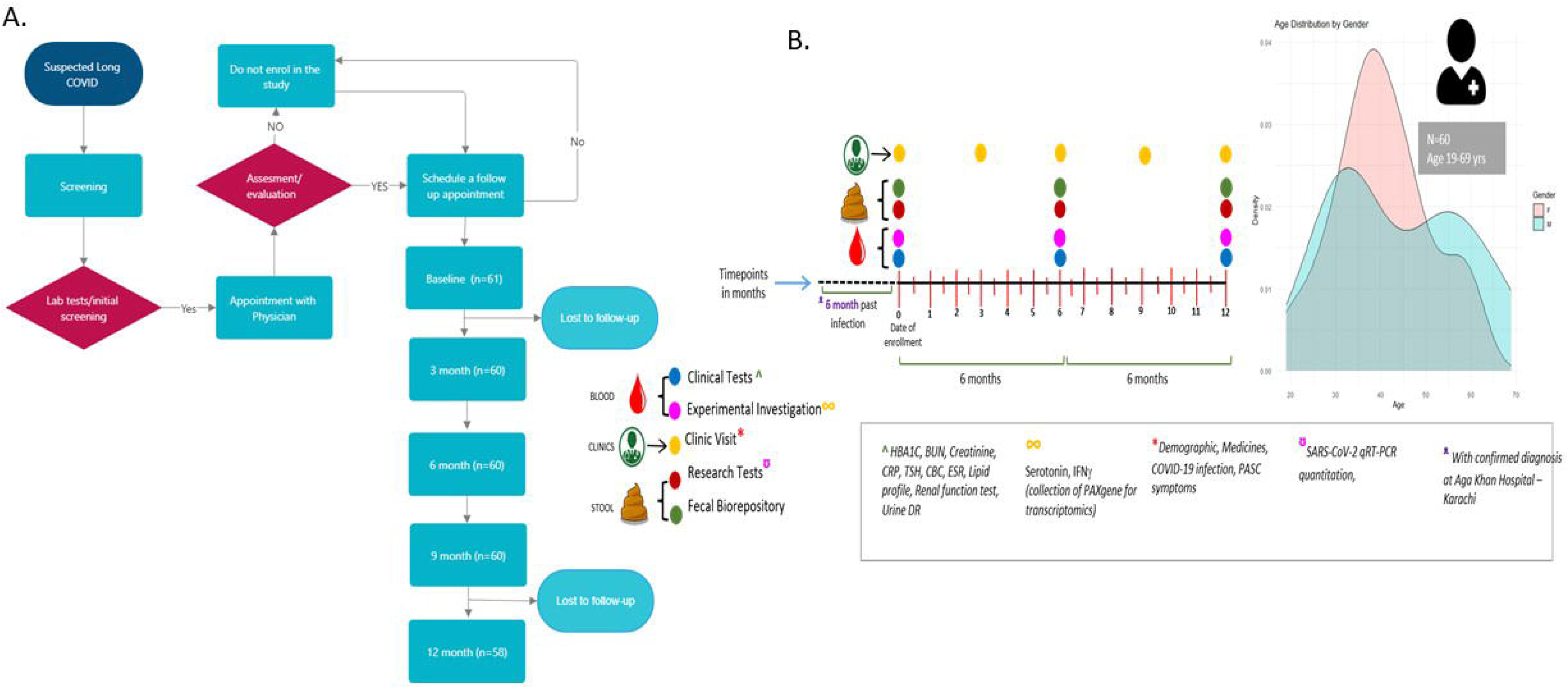
Study flow for enrolment, clinical and biological sample collection of PASC subjects. Figure 1A. shows a longitudinal study design with follow-up at the interval of 3 months for clinical follow-up (0,3,6,9&12 months) while biological sample (blood and Fecal) follow-up at 3 time points (0, 6 & 12 months), Figure 1B shows biological sample collection over the period of one year with clinical visits, clinical tests and research investigations

### RNA Extraction

RNA was extracted from the stool samples. Briefly, stool samples were thawed and vortexed thoroughly, then centrifuged to pellet down the solid particles. Clear supernatant of 140 µL was collected and processed using QIAamp® Viral RNA Extraction Kit (Cat # 52906, Qiagen-GmbH, Hilden, Germany) following the manufacturer’s protocol. The RNA was eluted in 80 μL of elution buffer and stored at –80°C.

### q-RT-PCR

Extracted RNA from faecal samples was subjected to RT-PCR for the detection of SARS-CoV-2 RNA using 2019-nCoV-PCR Mix and 2019-nCoV-PCR-Enzyme Mix (Cat #S3102ESC2) targeting the Orf1ab and N genes of the novel coronavirus (2019-nCoV). RT-PCR was performed using the Bio-Rad CFX96 instrument. The cycling conditions included an initial reverse transcription step at 50°C for 30 minutes, followed by cDNA pre-denaturation at 95°C for 1 minute, denaturation at 95°C for 15 seconds, followed by annealing, extension, and fluorescence collection at 60°C for 30 seconds (45 cycles) followed by device cooling to 25°C for 10 seconds. Fluorescence signals were detected using the FAM channel for the Orf-1ab gene and ROX channel for N gene. Any amplification curve crossing the threshold before 40 cycles was considered positive. Faecal shedding was assessed based on positivity in PCR for either the N or Orf-1ab gene.

### Serotonin ELISA

Serotonin ELISA was performed by using an ultrasensitive enzyme immunoassay for the quantitative determination of serotonin. Plasma samples were prepared by adding 800 µl of normal saline into 200 µl of plasma sample and centrifuged at 4500 g for 10 minutes at 4°C. After discarding the supernatant, the pellet was resuspended in 200 µl of ultrapure water and vortexed thoroughly. Diluent was prepared to a concentration of 1X, and samples were diluted to 1:100 and 1:1000. Standards, control and samples were added into respective wells of acyl plate followed by 25 µl of acyl buffer to all the wells. The plate was acylated for 30 minutes at room temperature (20-25°C) on a shaker (600rpm). Subsequently, 100 µl of the acylated standards, controls and samples were placed in the Serotonin 5HIAA microtiter strips. 25 µl of SER-AS was added to all wells, and the plate was incubated for 15-20 hours at 2-8°C. After incubation and washing, 100 µl of conjugate was added into all the wells and incubated for 30 minutes at room temperature on a shaker. 100 µl of substrate was added in all wells and incubated at room temperature for 20-30 minutes. Absorbance was read at 450 nm after placing 100 µl of stop solution in all the wells. The serotonin concentration was calculated using a standard curve and multiplied by a correction factor of 100 as per manufacturer’s instructions.

## Results

Most participants manifested long-term symptom persistence from the “days of illness during acute COVID-19” and “PASC symptom appearance”. We also noticed multiple re-infections of COVID-19 in participants. 17/60 patients had their first COVID-19 positive in 2020; of those, 9/17 had a second infection within (2021–2024) 1-4 years. 23/60 patients had a first infection in 2021, 4/23 had a second infection from 2022 to 2023, while 20/60 had a first infection in 2022 and 4/20 had a second infection within 3-6 months.

### Baseline characteristics

A total of 61 participants who fulfilled the study criteria were recruited from Aga Khan University; clinical assessment was done in the study Physician’s clinic through a structured questionnaire and clinical judgment to qualify as PASC subjects. PASC participants were interviewed by study physicians and enrolled in the study (Figure 1). Of these, one participant was lost to follow-up at the 3-month time point and was excluded from the study. Therefore, a total of 60 participants were included in the final analysis. Two participants were lost to follow-up at the 12-month time point; however, data from 60 participants were included in the analyses.

The primary outcome of the study was the presence and severity of PASC-related symptoms over time in the population of Pakistan. The secondary outcomes included longitudinal changes in clinical symptomatology associated with PASC-related symptoms over a period of 12 months.

The frequency of each symptom was analysed, and only symptoms with a frequency greater than 30% were selected for further analysis. A total of 22 symptoms met this criterion and were included in the analysis (Suppl. Figure S1).

Based on the selected 22 symptoms, a score was assigned to each participant at each time point, with the presence of a symptom corresponding to a score of 1. The maximum possible score at a single time point was 22, and the minimum was 0. At each time point, individual participant scores were used to calculate the median and standard deviation for the cohort. The median score was then assessed to evaluate trends and variations across different time points.

Sixty PASC subjects were enrolled from 2023-2024 in this study. Table 1 shows characteristics of the study group. The mean age ± SD of the study subjects was 42.1 ± 12.1 years, and 60.0% of the participants were female. The median BMI ± IQR for the sample population was 26.6 kg/m² (IQR 23.8-31.6); 20 participants (33.3%) had normal weight, 18 (30.0%) were overweight, and 22 (36.7%) were classified as having obesity (Class I-III). Vital parameters were largely within the normal range at baseline. All participants had confirmed SARS-CoV-2 infection; 43 (71.7%) reported one infection, and 17 (28.3%) reported multiple infections. Six participants (10.0%) had been hospitalized for COVID-19, 28 (46.7%) had at least one comorbidity, 30 (50.0%) had received three vaccine doses, and 7 (11.7%) reported tobacco use.

**Table 1:** Baseline demographics, clinical characteristics, and covid-19 history of the study population.

| **Variable** | **Overall (N=60)** |
| --- | --- |
| **Age (years)** | 42.1 (12.1) |
| **Sex at birth** |  |
| Female | 36 (60.0%) |
| Male | 24 (40.0%) |
| **BMI (kg/m<sup>2</sup>)** | 26.6 (23.8, 31.6) |
| **BMI categories** |  |
| Normal | 20 (33.3%) |
| Overweight | 18 (30.0%) |
| Class I obesity | 16 (26.7%) |
| Class II obesity | 4 (6.7%) |
| Class III obesity | 2 (3.3%) |
| **Vital signs** |  |
| Pulse rate (bpm) | 83.0 (74.0, 92.3) |
| Respiratory rate (breaths/min) | 18.5 (18.0, 20.0) |
| Body temperature (°C) | 36.2 (36.0, 36.4) |
| Systolic BP (mmHg) | 125 (120, 136) |
| Diastolic BP (mmHg) | 76.0 (68.0, 83.0) |
| Oxygen saturation (%) | 98.0 (98.0, 98.3) |
| Pain score (0–10) | 4.0 (0, 6.3) |
| **COVID-19 history** |  |
| Ever tested positive | 60 (100%) |
| Number of infections |  |
| 1 | 43 (71.7%) |
| 2 | 14 (23.3%) |
| 3 | 3 (5.0%) |
| Hospitalisation due to COVID-19 |  |
| No | 54 (90.0%) |
| Yes | 6 (10.0%) |
| **Comorbidities** |  |
| No | 32 (53.3%) |
| Yes | 28 (46.7%) |
| **COVID-19 vaccine doses** |  |
| 0 | 2 (3.3%) |
| 1 | 4 (6.7%) |
| 2 | 6 (10.0%) |
| 3 | 30 (50.0%) |
| 4 | 15 (25.0%) |
| 5 | 3 (5.0%) |
| **Tobacco use** |  |
| No | 53 (88.3%) |
| Yes | 7 (11.7%) |
| **Smoking status** |  |
| No | 54 (90.0%) |
| Yes | 6 (10.0%) |

### Changes in symptom scores over time

Supplementary Figure 2 shows the error bars for Scores of symptoms, which showed a steady decrease throughout the follow-up period. The average trend revealed a relatively slight drop from baseline to 3 months, after which there was an initial stabilization period at 6 months and a steeper decline at 9 and 12 months. The overall trend suggested continual improvement in symptoms with individual variations noted.

The results of the symptom scores showed a gradual decreasing trend in the follow-up period. To check whether the difference in score from the preceding time point is significant, we checked the difference from baseline and the preceding follow-up month. Table 2 shows the difference in score was observed for baseline vs. 3 months (p=0.063) and baseline vs. 6 months (p=0.082). However, compared to baseline, a significant decline in symptoms was observed at 9 months (p=0.001) and a further decrease at 12 months (p<0.001). While comparing the 3^rd^ month as the index for subsequent follow-ups, there was no significant difference observed except for 3 months vs. 12 months (p<0.001). With reference to 6 months, there was a significant decline observed at both 9 and 12 months (p<0.001). Moreover, there was no significant difference between 9 months and 12 months (p=0.524).

**Table 2.** Pairwise comparisons of symptoms scores at different time points.

| Group 1 | Median [Q1, Q3] | Group 2 | Median [Q1, Q3] | p-value |
| --- | --- | --- | --- | --- |
| Baseline | 12.0 [14.3, 6.0] | 3<sup>rd</sup> month | 9.0 [12.3, 6.0] | 0.063 |
| Baseline | 12.0 [14.3, 6.0] | 6<sup>th</sup> month | 9.0 [12.0, 7.0] | 0.082 |
| Baseline | 12.0 [14.3, 6.0] | 9<sup>th</sup> month | 8.0 [10.0, 6.0] | **0.001** |
| Baseline | 12.0 [14.3, 6.0] | 12<sup>th</sup> month | 7.0 [11.0, 4.0] | **<0.001** |
| 3<sup>rd</sup> month | 9.0 [12.3, 6.0] | 6<sup>th</sup> month | 9.0 [12.0, 7.0] | 0.844 |
| 3<sup>rd</sup> month | 9.0 [12.3, 6.0] | 9<sup>th</sup> month | 8.0 [10.0, 6.0] | **0.002** |
| 3<sup>rd</sup> month | 9.0 [12.3, 6.0] | 12<sup>th</sup> month | 7.0 [11.0, 4.0] | **<0.001** |
| 6<sup>th</sup> month | 9.0 [12.0, 7.0] | 9<sup>th</sup> month | 8.0 [10.0, 6.0] | **<0.001** |
| 6<sup>th</sup> month | 9.0 [12.0, 7.0] | 12<sup>th</sup> month | 7.0 [11.0, 4.0] | **<0.001** |
| 9<sup>th</sup> month | 8.0 [10.0, 6.0] | 12<sup>th</sup> month | 7.0 [11.0, 4.0] | 0.524 |

**Footnote:** p-value computed using Wilcoxon Signed-Rank Test.

### Symptom pattern evolution

Figure 2 shows changes in the frequency distribution of symptoms using radar plot analysis. The symptoms are interpreted as 100% in the outer circle and 25% in the innermost circle. At the initial assessment stage or baseline, the frequency of symptoms was diverse, spanning a range of symptoms including musculoskeletal, neurological, and pain-related symptoms. The most significant symptoms were fatigue, myalgia, weakness, and breathlessness. At baseline, symptoms of “Fatigue” were (57/60) 95%, [3 months (50/60) 83%, 6 months (48/60) 80%, 9 months (50/60) 83%] which gradually decreased to 63% (38/60) at 12 months. Myalgia remained relatively static throughout, at 60% (36/60) at baseline [3 months (43/60) 72%, 6 months (37/60) 62%, 9 months (46/60) 78%], and (38/60) 63% at 12 months. However, symptoms of weakness were reduced from 76% (46/60) [3 months (45/60) 75%, 6 months (41/60) 68%, 9 months (47/60) 78%] to 58% (35/60) at 12 months. Similarly, breathlessness was improved from 63% (38/60) at baseline [3 months (25/60) 42%, 6 months (30/60) 50%, 9 months (17/60) 28%] to 32% (19/60) at 12 months. A few other symptoms showed dynamic reduction such as anosmia [baseline, 30% (18/60); 12 months 6.6 % (4/60)] brain fog [baseline, 33% (20/60); 12 months 21% (13/60)], recurrent fever [baseline, 45% (27/60); 12 months 12% (7/60)], and headache [baseline, 35% (21/60); 12 months 15% (9/60)]. This suggests an improvement in overall symptoms, with heterogeneity in the rate of symptom reduction. (Figure 2).

**Figure 2:**
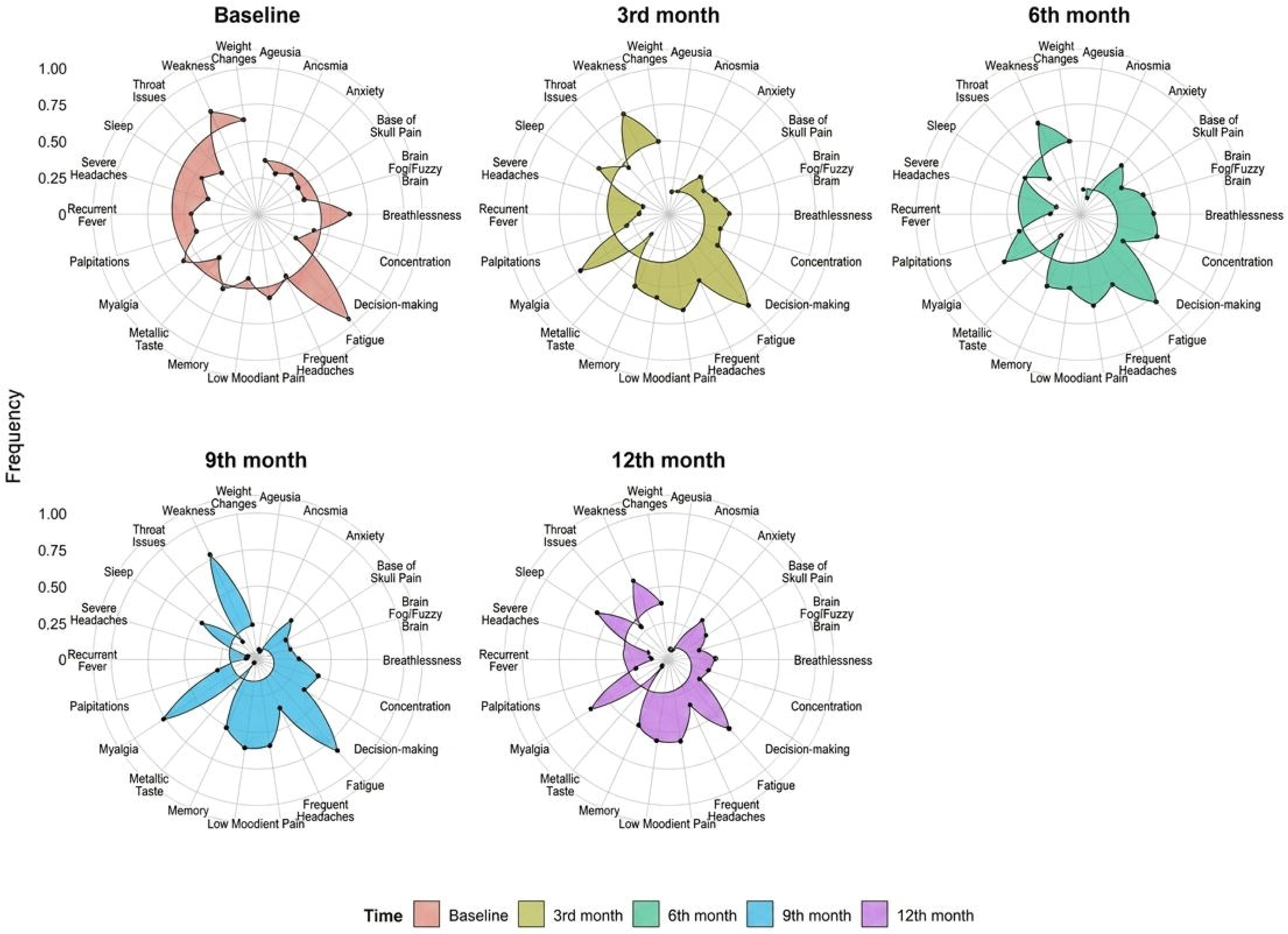
Symptoms of PASC captured over 12 months. Radar plots showing the distribution of frequencies across 22 symptoms at baseline and follow-up assessments (3, 6, 9, and 12 months). The symptoms are interpreted as 100% in the outer circle and 25% in the innermost circle. Each axis represents a domain, and the distance from the center indicates the frequency. Changes in the size and shape of the plots over time reflect temporal variation in domain!zlspecific patterns.

### Clustering of symptoms

Hierarchical clustering of the longitudinal symptom trajectories identified four distinct symptom groups (Figure 3). Weakness and fatigue formed a distinct cluster characterized by persistently high symptom prevalence throughout the 12-month follow-up. Sleep disturbance, low mood, memory impairment, myalgia, and joint pain formed a second distinct cluster with a gradual decline in prevalence over time. A third cluster consisting of symptoms of anosmia, ageusia, recurrent fever, metallic taste, and severe headaches demonstrated rapid resolution during follow-up. The last cluster had heterogeneous symptoms spanning neurocognitive and autonomic symptoms, including brain fog, concentration difficulties, decision-making impairment, anxiety, palpitations, breathlessness, throat symptoms, weight changes, pain in the base of the skull, and frequent headaches, which showed moderate persistence with slower recovery than the sensory symptoms (Figure 3). Figure 4 shows similar observations of symptoms recorded in a group-wise analysis. The frequency was maximum for Constitutional symptoms at 0 months, 69.6% (167/240), compared to 3 months, 57% (137/240), followed by 6 months, 54% (130/240), 9 months, 48% (115/240), and 12 months, 42.5% (102/240). Constitutional symptom categories showed a decreasing trend, whereas Neurocognitive [0M: 40% (97/240), 3M: 41.25% (99/240), 6M: 47.5%(114/240), 9M: 39.16% (94/240),12M: 31.25% (75/240)]; Musculoskeletal [0M: 59% (71/120), 3M: 69% (83/120),6M: 62.5% (75/120), 9M: 68% (82/120),12M: 60% (72/120)] and Psychological symptoms [0M: 40% (48/120),3M: 45.8% (55/120), 6M: 47.5% (57/120),9M: 48.3% (58/120),12M: 45.8% (55/120)] have persisted from baseline throughout 12 months follow-up with no significant change.

**Figure 3:**
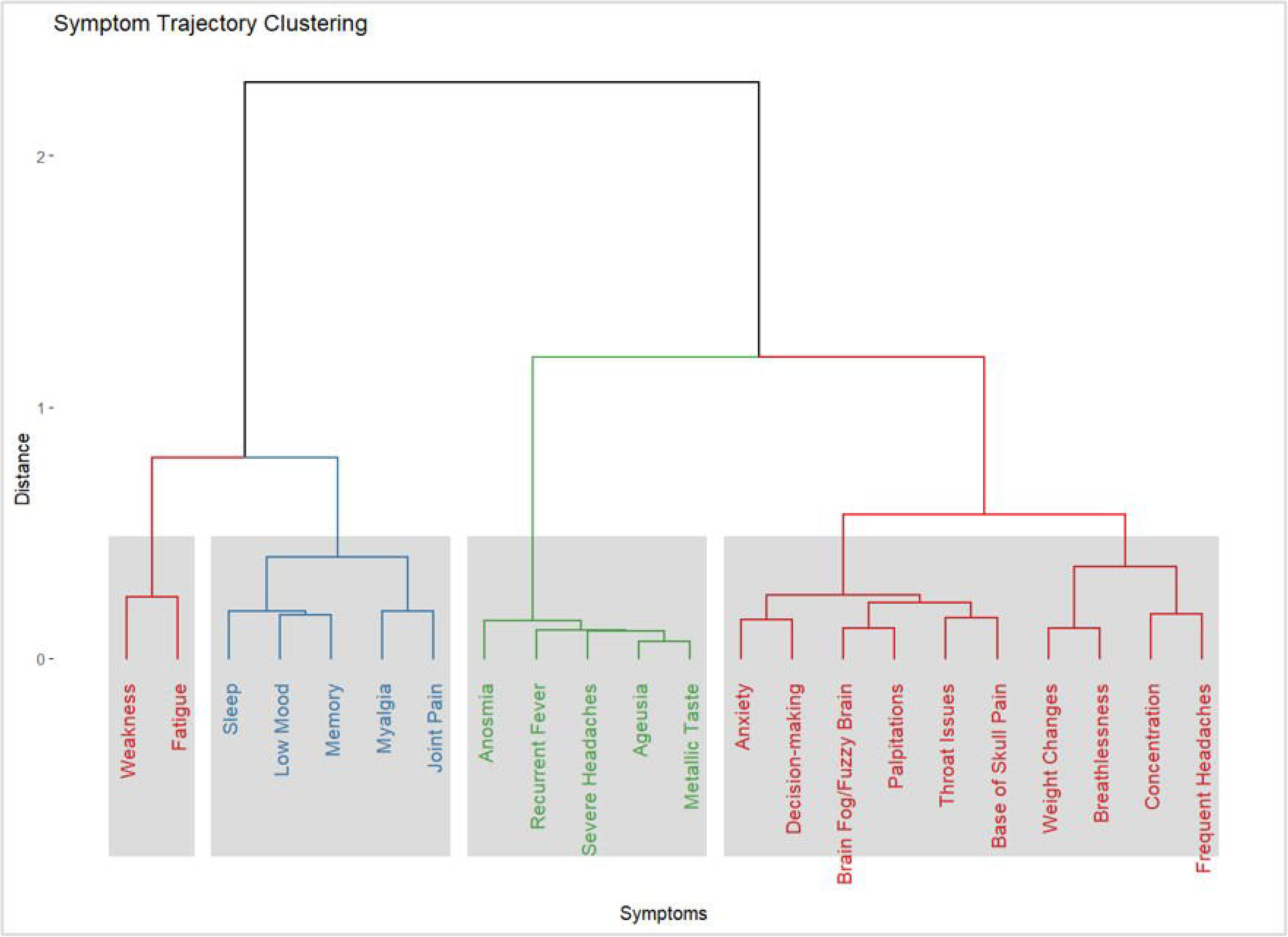
PASC Symptoms profiling at baseline. Hierarchical clustering dendrogram of symptom profiles, with the y!zlaxis representing dissimilarity. Distinct clusters of symptom patterns are highlighted by dashed boxes.

**Figure 4:**
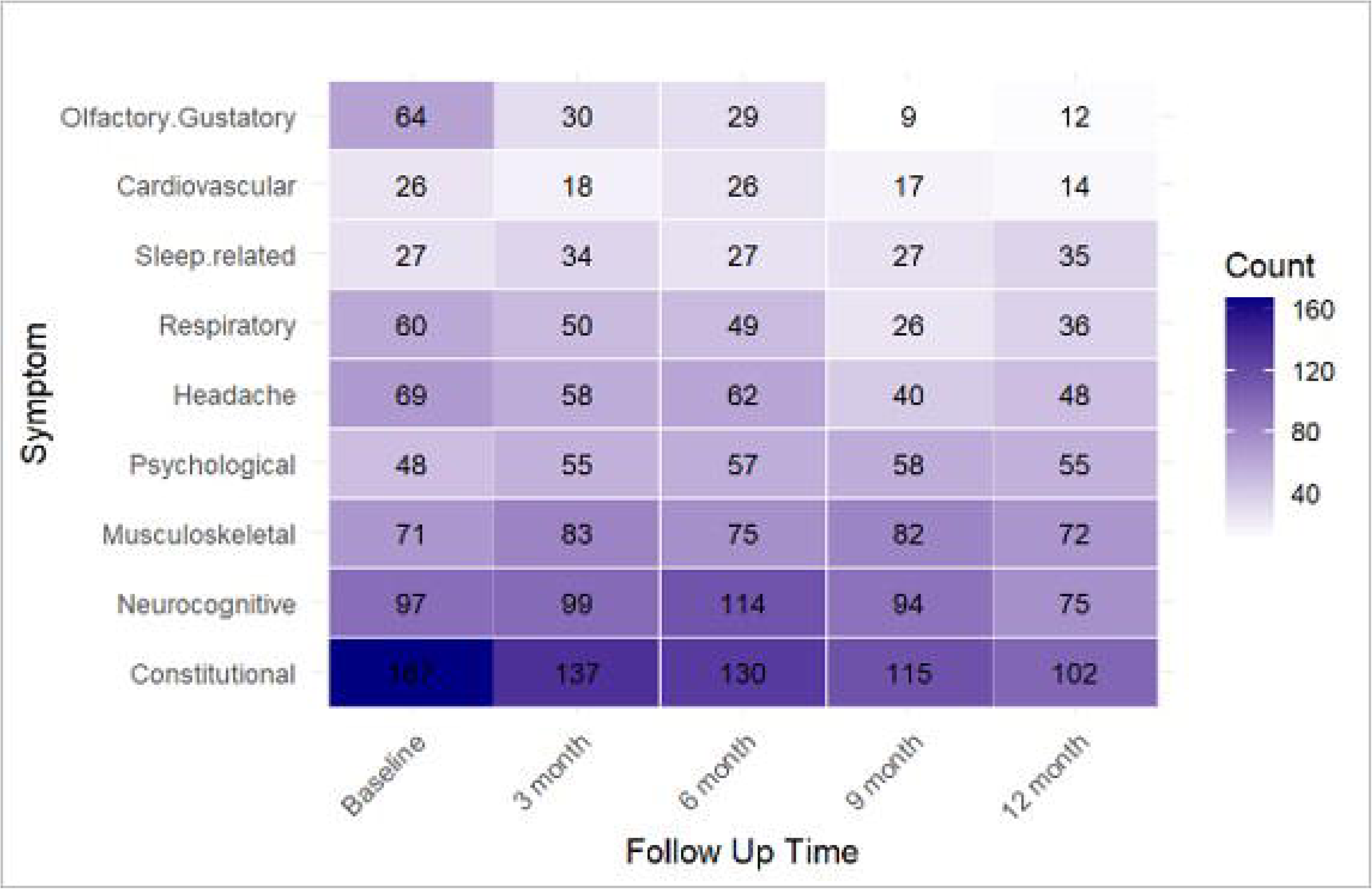
PASC symptoms frequency heatmap over 12 months. Heatmap shows group wise symptom frequencies distribution across follow!zlup time points (baseline, 3, 6, 9, and 12 months), with darker shading indicating higher frequency.

Suppl Fig 1 shows the heatmap of symptoms rank-ordered, with highest to lowest frequency as depicted by colour gradient. The frequency of symptoms was highest at the initial assessment, with fatigue, myalgia, weakness, joint pain, and sleep disturbances being the topmost symptoms. There was an overall decrease in the frequency of symptoms during the follow-up period; however, fatigue, myalgia, weakness, and joint pain were persistently reported by over half of the participants. The prominent decline was observed in sensory symptoms such as anosmia and ageusia within 3 to 6 months.

### Longitudinal mixed-effects model of symptom trajectories

Table 3 shows longitudinal assessment of data in a Linear Mixed Effects Model. There was a statistically significant decline in the symptom score throughout the study period. As compared to the baseline period, there was a decrease in the symptom score at 9 months (β = −2.68, 95% CI: −3.81 to −1.55; p < 0.001) and 12 months (β = −3.00, 95% CI: −4.13 to −1.87; p < 0.001). There was no statistically significant difference in estimates at 3 months (β = −1.08; p = 0.063) and 6 months (β = −1.00; p = 0.086). Overall, we found a strong negative trend over time (time effect: β = −0.25, 95% CI: −0.35 to −0.11; p < 0.001). For other covariates such as smoking positively associated with high symptom score (β = 5.13, 95% CI: 2.77 to 7.50; p < 0.001), Class III obesity group was also positively associated with a high symptom score (β = 7.27, 95% CI: 2.54 to 11.99; p = 0.014). There was no statistical significance for other covariates.

**Table 3.** Linear mixed-effects model estimating change in symptoms score.

| Variable | Estimate ($\beta$) | 95% CI | P-value |
| --- | --- | --- | --- |
| **Time (ref = Baseline)** |  |  |  |
| 3<sup>rd</sup> month | -1.08 | -2.21 - 0.05 | 0.063 |
| 6<sup>th</sup> month | -1.00 | -2.13 - 0.13 | 0.086 |
| 9<sup>th</sup> month | -2.68 | -3.81 - -1.55 | **<0.001** |
| 12<sup>th</sup> month | -3.00 | -4.13 - -1.87 | **<0.001** |
| **Time (Overall)** | -0.25 | -0.35 - -0.11 | **<0.001** |
| **Age (>40 vs ≤40)** | 1.20 | -0.39 - 2.80 | 0.215 |
| **Gender (Male vs Female)** | -1.65 | -3.31 - 0.01 | 0.106 |
| **BMI (ref = Normal)** |  |  |  |
| Class I Obese | -0.18 | -2.02 - 1.66 | 0.869 |
| Class II Obese | 1.31 | -1.70 - 4.31 | 0.472 |
| Class III Obese | 7.27 | 2.54 - 11.99 | **0.014** |
| Overweight | 0.66 | -1.15 - 2.46 | 0.549 |
| **COVID frequency (ref = One)** |  |  |  |
| Two times | 0.94 | -0.87 - 2.75 | 0.392 |
| Three times | 1.42 | -2.02 - 4.85 | 0.495 |
| **Hospitalised (Yes vs No)** | -0.71 | -3.19 - 1.77 | 0.636 |
| **Comorbidities (Yes vs No)** | 1.66 | 0.02 - 3.30 | 0.099 |
| **Vaccine doses (ref = 0)** |  |  |  |
| 1 dose | 1.01 | -4.56 - 6.58 | 0.764 |
| 2 doses | 0.25 | -5.18 - 5.68 | 0.94 |
| 3 doses | 1.51 | -3.34 - 6.35 | 0.608 |
| 4 doses | 1.02 | -4.03 - 6.07 | 0.738 |
| **5 doses** | 6.78 | 0.56 - 13.01 | 0.077 |
| **Days since infection** | -0.00059 | -0.0032 - 0.0021 | 0.714 |
| **Smoking (Yes vs No)** | 5.13 | 2.77 - 7.50 | **<0.001** |

**Footnote:** Estimates ( $\beta$ ) are from a linear mixed-effects model. All results are adjusted for covariates listed in the table. Time is modelled with baseline as reference. $\beta$ represents change in symptom score; p < 0.05 indicates statistical significance.

### Viral faecal shedding

We also analysed longitudinal SARS-CoV-2 faecal shedding in PASC subjects for viral Nucleocapsid (N) and Open Reading Frame (orf-1ab) gene at baseline, 6 months, and 12 months. We identified three types of faecal shedders in our cohort.

**Baseline shedders (BS)** defined as individuals who were positive for shedding at baseline but became negative at both the 6-month and 12-month follow-up assessments.

**Persistent shedders (PeS)** were those who remained positive for shedding from baseline through the 6-month and/or 12-month time points, indicating a continuous or sustained pattern of shedding over time.

**Intermittent shedders (IS)** were identified as individuals who were negative for shedding at baseline but exhibited positive results at either the 6-month or 12-month follow-up, or who were positive at baseline but exhibited negative results at 6-month but were positive again at 12-month, suggesting episodic or reactivated shedding.

In summary, we had **4** Intermittent shedders (IS) who were negative at baseline, **7** persistent shedders (PeS) who were positive at baseline as well as 6 months, and **23** Baseline (BS) shedders who were positive at baseline only.

Figure 5 shows a density plot of Ct values of all three types of faecal shedders, which range from 31.02 to 39.78 for N gene, while the Orf-1ab gene ranges from 30.52 to 44.28. The IS had a Ct value ranging from 34 to 37, PeS had 32.54 to 38.07, and BS had a Ct value ranging from 31.02 to 39.78.

**Figure 5:**
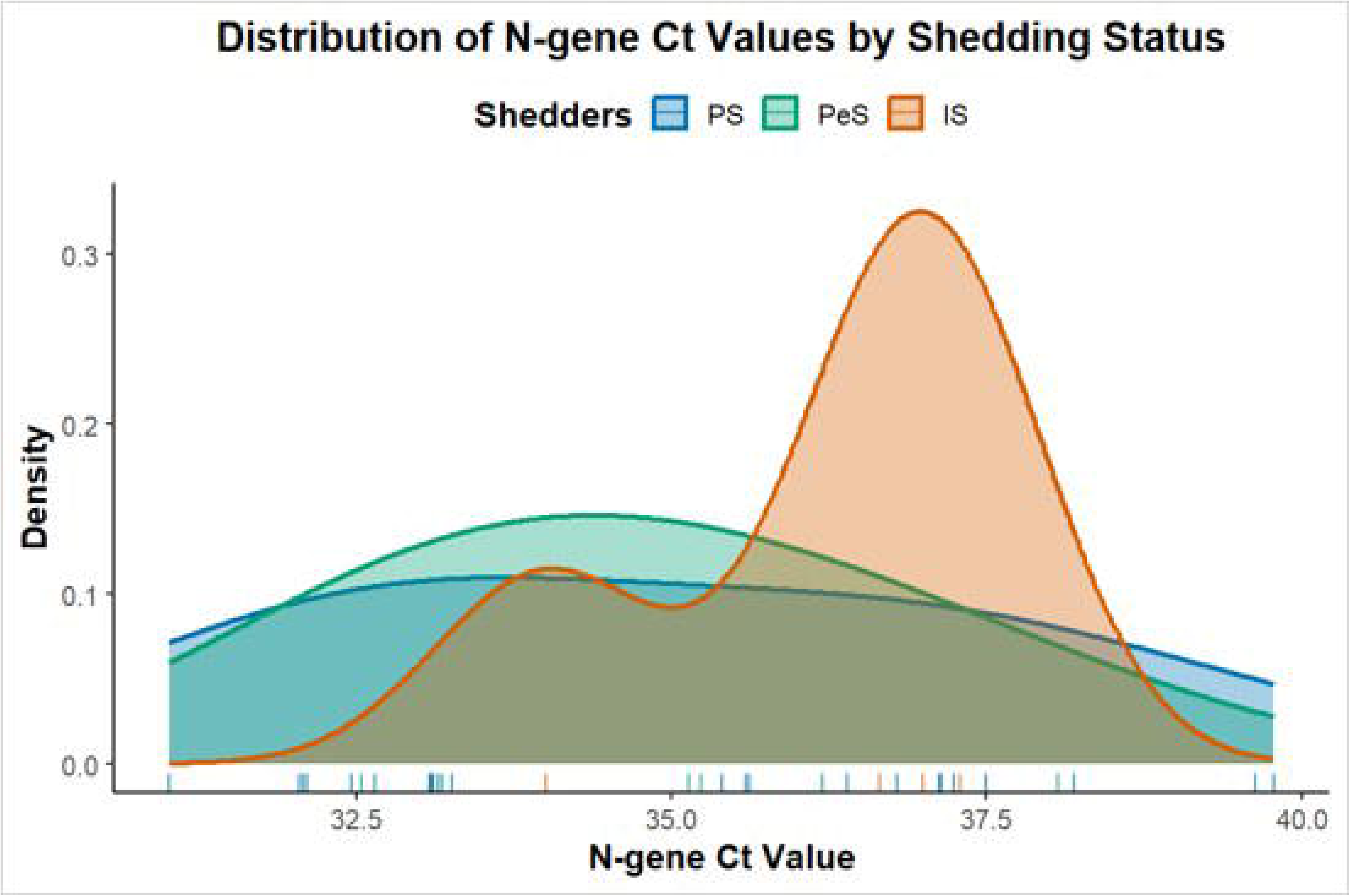
Quantitative Ct value of viral faecal shedding in PASC. Viral faecal shedding in Baseline shedder (BS), Persistent Shedder (PeS) and Intermittent Shedder (IS). The X axis shows Ct values and Y axis shows density of Ct values for categories (IS, BS, PeS).

Next, we assessed the presence of symptoms in the faecal shedder vs. non-shedders at baseline and 6 months (Suppl Table 2). Of all 22 symptoms at baseline, joint pain (63%) was more common in shedders compared to non-shedders, while other symptoms were more prevalent in non-shedders, such as Fatigue (96.7%), Weakness (80%), Low mood (60%), Myalgia (66.7%), and Memory (66.7%). Compared to the baseline, at 6 months there was an overall increase in symptom frequency in shedders such as Breathlessness (77.8%), Joint pain (88.9%), Frequent headaches (55.6%), Anxiety (55.6%) and Weakness (77.8%), metallic taste (22.2%) (Figure 6).

**Figure 6:**
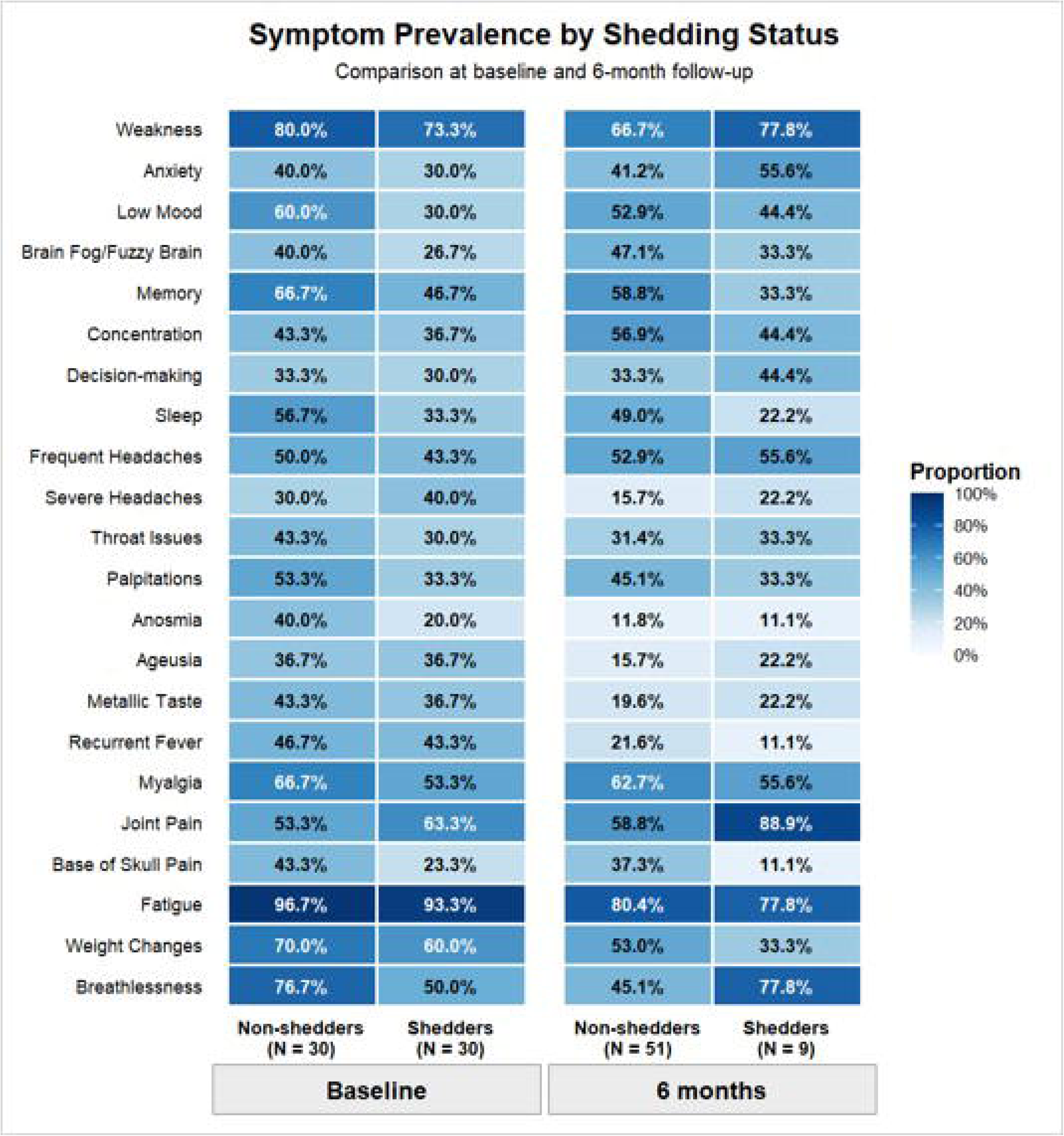
PASC symptoms frequency heatmap in Shedders and Non-shedders. Heatmap of symptoms and with faecal shedding status at baseline and 6 months.

### Serotonin analyses

We next analysed serotonin levels in platelet-rich plasma samples in Acute COVID-19 (n=77), and PASC subjects (n=59) at baseline, 6 months, and 12 months. Figure 7 compares the median-IQR levels of Serotonin in PASC subjects at baseline (0.68; 0.10-9.61), 6 months (1.48; 0.65-3.83), and 12 months (0.92; 0.03-2.21) compared to Acute COVID-19 cases (0.60; 0.60-0.77). There was no significant difference in serotonin levels between the acute phase of COVID-19 and the sequelae of Long COVID. There was a trend of decreasing serotonin in PASC subjects after 6 months, with a significant decrease observed between 6 and 12 months (p=0.02).

**Figure 7:**
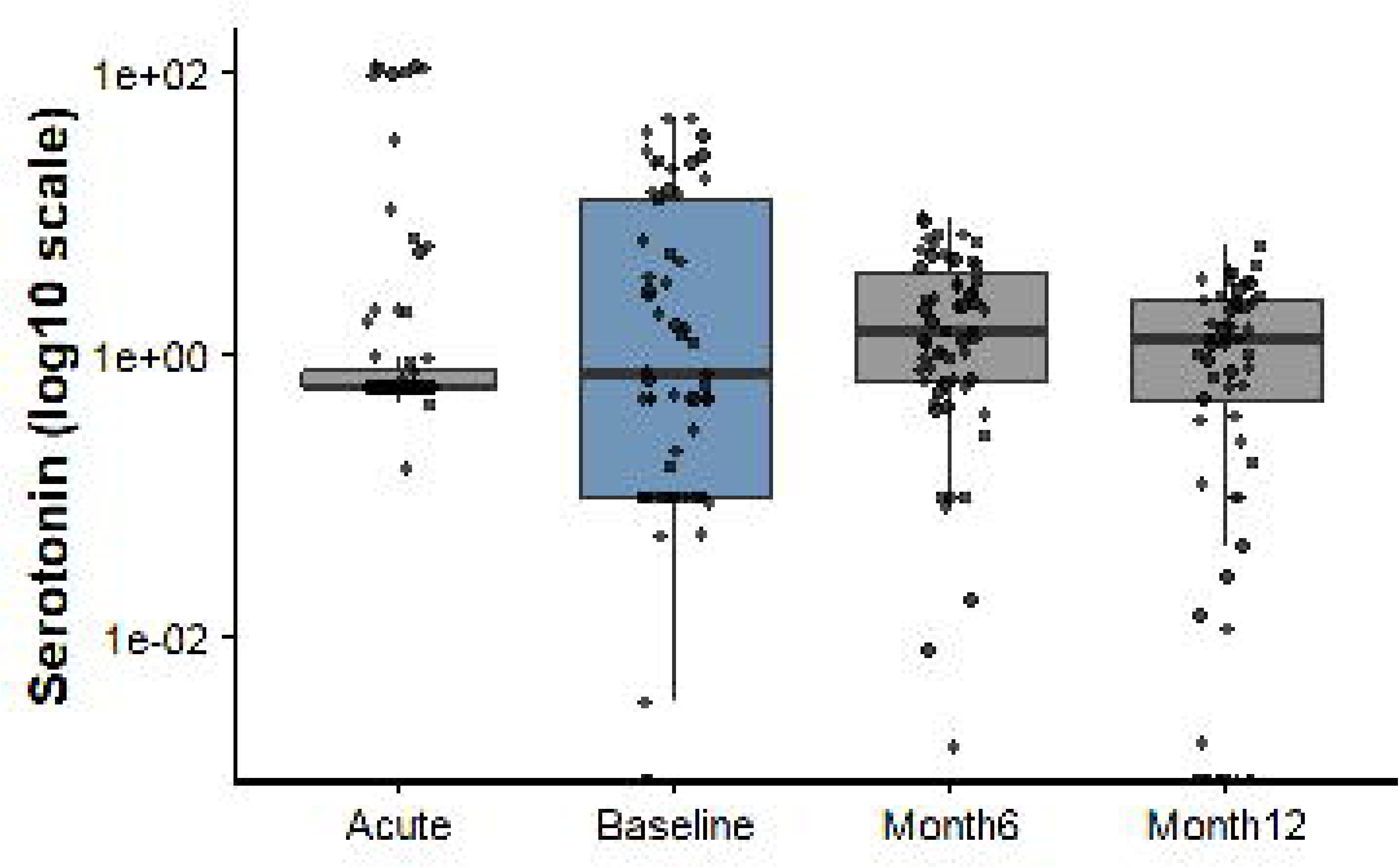
Estimation of Serotonin in PASC and Acute COVID-19 patients. Serotonin level in Acute COVID-19 (n=77), and PASC subjects at baseline (n=59), 6 months (n=59), and 12 months (n= 59). Serotonin level was measured by sandwich ELISA in platelet-rich plasma (PRP). Briefly PRP was incubated in acylated plates for 16-20 hrs at 4 LJC. The samples were transferred to serotonin 5-HIAA plates and incubated for 2hrs. Finally, the plates were read at 450 nm. The median values were compared using the Kruskal-Wallis test (p=0.04). The post hoc test was significant only for median values of 6 and 12 months (p=0.02).

Comparing Serotonin level with viral faecal shedding, we observed viral persistence at baseline (n=30), 6 months (n=9), and 12 months (n=4), which was consistent with serotonin levels observed in PASC subjects at baseline (Median = 0.6 pg./ml), which was increased at 6 month (median =1.48 pg./ml), and dropped at 12 month (Median =0.92 pg./ml), consistent with a marked reduction in faecal shedding at 6 (n= 9), and 12 (n=4) months.

## Discussion

This is the first report of a longitudinal cohort on clinical phenotypes of PASC from a tertiary care setting in Pakistan. This study reports a combination of symptom recording through a structured survey and lab biomarker assessment over a period of one year. PASC has invariably affected many populations, but data from LMICs are restricted, with a few reports from India(23), Bangladesh(25) and Pakistan(21). In an analysis of 500 studies, 90% of studies represented data from high-income settings (26). This shows the relative lack of representation of Southeast Asian cohorts in understanding the burden and clinical phenotypes of PASC in this region.

There was a high prevalence of persistent post-COVID-19 symptoms among females in our cohort that was consistent with findings from several previous studies(27, 28). This might be related to hormonal factors, predominantly oestrogen-mediated immune inflammation, or genetic predisposition to autoimmune diseases, and prolonged/sustained immune dysregulations commonly seen in females (28, 29).

Our study suggests that the development and persistence of PASC symptoms may occur independent of the severity of the initial acute SARS-CoV-2 infection, as most patients were not hospitalized and had mild infection. Consequently, individuals with mild or asymptomatic disease can experience long-term post-COVID sequelae comparable to those observed following severe illness(30). This is in accordance with our study, where only 10% of patients who developed symptoms of PASC had severe or critical COVID-19 (31).

Uncontrolled diabetes is associated with a high risk of development of PASC syndrome(32), especially cognitive impairment and respiratory complications. Most of our study subjects had Type II diabetes mellitus, and 55% of participants had higher FBS levels (>100mg/dL). Similarly, comorbid conditions (diabetes and hypertension) and metabolic syndrome are well-established risk factors for dyspnea, cognitive impairment, and fatigue, which involve pathways of persistent inflammation, immune dysregulation (17), endothelial dysfunction, activation of the renin–angiotensin–aldosterone system (RAAS), oxidative stress (33, 34) (35, 36) and metabolic disturbances (37).

Stringent glycaemic control, blood pressure management, and weight reduction can be targeted in the high-risk group to mitigate the development of PASC.

Overall, we found a substantial burden of constitutional symptoms at baseline. Fatigue, myalgia, and breathlessness were the most common symptoms, with other neurological symptoms (38), such as cognitive difficulties, low mood, difficulties in decision making, anxiety, sleep disturbances, and brain fogginess, as being the most prevalent manifestations among PASC subjects (39). The prevalence of PASC symptoms continued to decline at 12 months; however, fatigue, musculoskeletal/arthralgia, and psychological dysfunction persisted in subjects, highlighting the prolonged burden of PASC. A range of 20-25% of acute COVID-19 patients report persistence of at least one symptom beyond 1 year (40, 41). Persistent symptoms might be attributed to ongoing inflammation and immune dysregulation of the immune system (4, 42).

Fatigue remained the predominant PASC symptom over the 12-month follow-up period. Although its frequency declined over time, more than one-third of patients continued to experience fatigue up to 1-year follow-up, underscoring its persistence and potential impact on functional recovery and quality of life. This finding is consistent with previous studies identifying fatigue as one of the most common manifestations of post-COVID syndrome(24, 43, 44).

Multisystem involvement is driven by a complex interconnection of organ systems(8), probably due to widespread distribution of ACE2 receptors in various organs, complement pathway dysregulation(45), viral infiltration of tissues, autoantibody generation, endothelial injury, and microvascular thrombosis(46). These factors collectively contribute to a chronic, self-perpetuating inflammatory milieu that persists long after the acute viraemic phase (47–49).

A prospective observational study showed an increased prevalence of inflammatory biomarkers and autoantibodies, particularly ANA, with higher titres correlated with persistent and severe fatigue (44). Multiple therapeutic interventions, such as herbal medicine, hyperbaric oxygen therapy, and structured rehabilitation programs, have shown potential benefit in improving symptoms of fatigue, physical performance, respiratory function, and quality of life in patients with post-COVID fatigue (50), with regular pacing, rehabilitation, and adoption of techniques to improve sleep quality (51–53).

Musculoskeletal symptoms (myalgia, arthralgia) were another group of persistent symptoms observed at 12 months. The prevalence of arthralgia was found to range from 2% to 65% between 4 months and 12 months, with a wide spectrum of manifestations such as arthralgia, fibromyalgia, and inflammatory arthritis, as new-onset or worsening pre-existing rheumatic conditions (54–56).

Among the various clinical manifestations, cognitive dysfunction is one of the most frequent manifestations of PASC, which is characterised by impaired concentration and decision-making ability, memory deficits, slowed thinking, and reduced mental clarity. This has substantially affected the daily functioning and quality of life of patients. Middle age, female gender, high BMI, and presence of comorbid illnesses were reported as risk factors associated with post-COVID cognitive dysfunction (57, 58). Although the precise pathogenesis is unclear, several possible mechanisms may be responsible for the pathophysiology of post-COVID cognitive dysfunction (59). These include persistent inflammation, latent viral reactivation, hypoxia, vascular damage, and possible direct viral invasion of the central nervous system (60, 61).

A few targeted therapeutic interventions and the timely implementation of cognitive rehabilitation, behavioural therapy, non-invasive brain stimulation, and hyperbaric oxygen therapy can lead to a reduction in symptoms and improvement in quality of life (62, 63).

The most biologically plausible mechanism underlying the symptoms of post-acute sequelae of SARS-CoV-2 infection (PASC) is the persistence of the virus or viral remnants within the body. This hypothesis is supported by studies demonstrating prolonged fecal shedding of SARS-CoV-2 (9) and its association with gastrointestinal symptoms (63). The persistence of viral particles and spike protein may lead to continuous immune stimulation, potentially contributing to the immune dysregulation (17, 49) observed in many individuals with Long COVID. An interesting observation in our study was the intermittent shedding detected in some patients, which appeared to coincide with fluctuations in their symptoms. This finding may indicate either viral persistence within the gastrointestinal tract (64) or repeated environmental exposure to the virus through potential oro-fecal transmission. A limitation of our study is that longer follow-up sampling was not feasible because of poor patient compliance. We did not observe any correlation between fecal shedding and symptoms at baseline. Additionally, the case report form (CRF) was not specifically designed to capture key gastrointestinal symptoms, which may have limited our ability to identify associations between gastrointestinal manifestations and fecal shedding.

Building on the findings related to fecal shedding, we also assessed serotonin levels in subjects with PASC and in individuals with acute COVID-19 (samples archived in repository). Viral persistence in the gastrointestinal tract is thought to be linked with continuous depletion of circulating serotonin through activation of Type I Interferon in the acute phase of COVID-19 and continues during PASC. In a UPenn PASC cohort, serotonin levels (5-HT) were significantly reduced in PASC with a concomitant decrease in metabolites, most importantly Tryptophan (65). Serotonin is produced from enterochromaffin cells in the intestine and stored in platelets. Two pathways are mainly involved in Serotonin depletion : thrombocytopenia, hypercoagulability, or IFNα activation by virus (65). This process may subsequently disrupt vagal nerve signalling, compromise the gut-brain axis and contribute to symptoms such as depression, low mood, and cognitive impairment (“brain fog”).

Our results demonstrated a decreasing trend in serotonin levels over time, with a significant decrease in serotonin levels at 12 months after infection, which is consistent with symptom resolution and the clearance of viral remnants from the body. Correspondingly, the proportion of subjects reporting brain fog and concentration difficulties decreased during follow-up. In contrast, symptoms such as low mood and sleep disturbances showed little to no improvement over the same period.

In conclusion, this study represents the first longitudinal investigation of PASC (Long COVID) in Pakistan, comprehensively evaluating multiple symptom domains, including constitutional symptoms (fatigue, weakness, and weight changes), neurocognitive, musculoskeletal, cardiovascular, and psychological manifestations. Our findings demonstrate that these symptoms can persist for years following acute SARS-CoV-2 infection, highlighting a significant burden on health and impacting wellbeing of individuals. Although a gradual decline in symptom frequency and severity was observed over time, complete resolution was not achieved in a few participants.

We also identified persistent faecal shedding of SARS-CoV-2 in a subset of our cohort, a finding that has not previously been reported from Pakistan. While faecal positivity was associated with modest fluctuations in symptom profiles, it did not appear to have a substantial impact on the overall burden of symptoms. These findings support the hypothesis of viral persistence and warrant further investigation into its role in the pathophysiology of Long COVID.

Future research should also examine the long-term sequelae of other viral infections in South Asian populations, who may be particularly susceptible to immune dysregulation and its associated health outcomes.

## Data Availability

All data produced in the present study are available upon reasonable request to the authors

