## Supplementary figures and images for "Clinical Phenotypes of Post-Acute Sequelae of SARS-CoV-2 (PASC) Infection: A Longitudinal Cohort Study from Karachi, Pakistan"

### Supplemental Figure S1

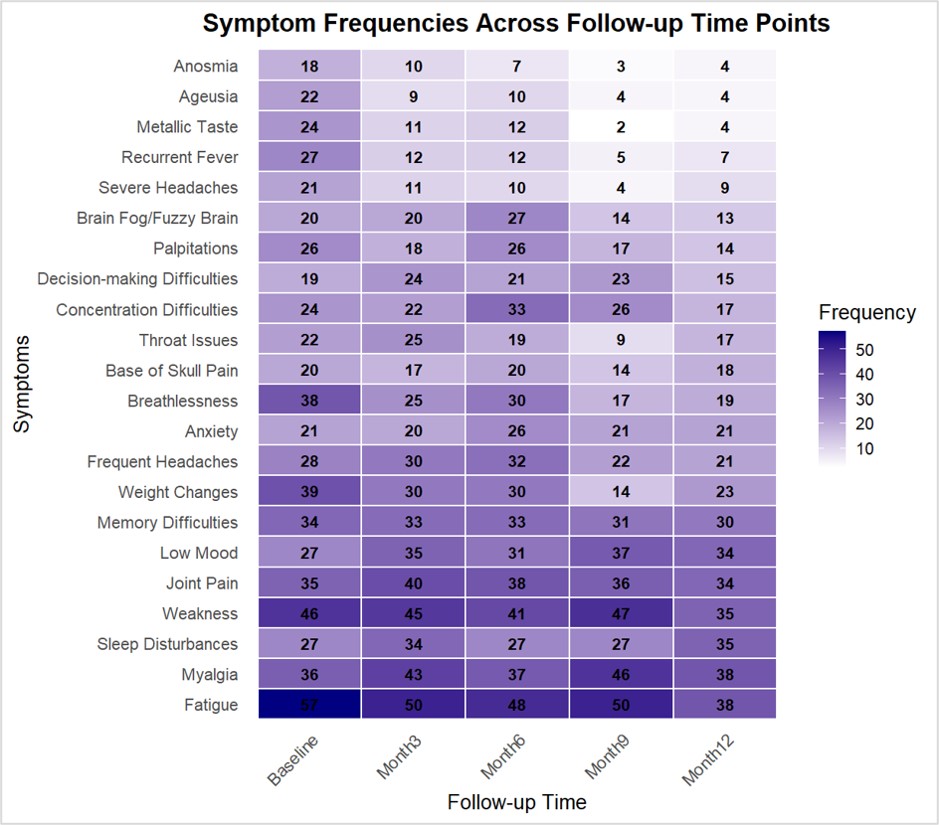

### Supplemental Figure S2

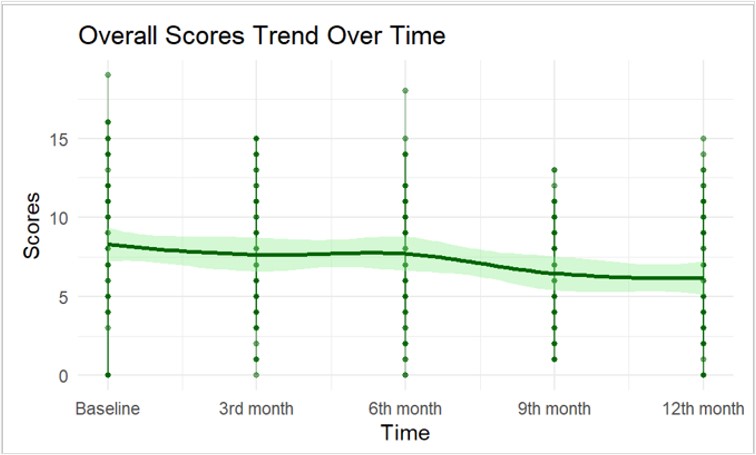
