## Supplementary Table 1 and 2 for "Clinical Phenotypes of Post-Acute Sequelae of SARS-CoV-2 (PASC) Infection: A Longitudinal Cohort Study from Karachi, Pakistan"

Suppl. Table 1: System-wise categorization of post-acute sequelae of SARS-CoV-2 (PASC) symptoms

|  | System | Symptoms | Frequency (%) |
| --- | --- | --- | --- |
| 1. | Constitutional | 1a.Fatigue | **57 (95.0%)** |
|  |  | 1b.Weakness | **46 (76.7%)** |
|  |  | 1c.Weight Changes | **39 (65.0%)** |
|  |  | 1d.Recurrent Fevers | **27 (45.0%)** |
| 2. | Neurocognitive | 2a.Brain Fog / Fuzzy Brain | 20 (33.3%) |
|  |  | 2b.Memory Difficulties | **34 (56.7%)** |
|  |  | 2c.Concentration Difficulties | **24 (40.0%)** |
|  |  | 2d.Decision-Making Difficulties | 19 (31.7%) |
| 3. | Psychological | 3a.Anxiety | **21 (35.0%)** |
|  |  | 3b.Low Mood | **27 (45.0%)** |
|  |  | 3c.Flashbacks | 17 (28.3%) |
| 4. | Sleep-related | 4a.Sleep Disturbances | **27 (45.0%)** |
|  |  | 4b.Nightmares | 7 (11.7%) |
| 5. | Headache | 5a.Frequent Headaches | **28 (46.7%)** |
|  |  | 5b.Severe headaches | **21 (35.0%)** |
|  |  | 5c.Base of Skull Pain | 20 (33.3%) |
| 6. | Neuro-ophthalmologic | 6a.Visual Disturbances | 17 (28.3%) |
| 7. | Respiratory | 7a.Cough | 17 (28.3%) |
|  |  | 7b.Breathlessness | **38 (63.3%)** |
|  |  | 7c.Nasal Issues | 15 (25.0%) |
|  |  | 7d.Voice Issues | 16 (26.7%) |
|  |  | 7eThroat Issues | **22 (36.7%)** |
|  |  | 7f.Throat Restriction | 13 (21.7%) |
| 8. | Cardiovascular | 8a.Chest Pain | 16 (26.7%) |
|  |  | 8b.Palpitations | **26 (43.3%)** |
| 9. | Olfactory/ Gustatory | 9a.Anosmia | 18 (30.0%) |
|  |  | 9b.Ageusia | **22 (36.7%)** |
|  |  | 9c.Metallic Taste | **24 (40.0%)** |
| 10. | Otolaryngological | 10a.Tinnitus | 17 (28.3%) |
| 11. | Musculoskeletal | 11a.Myalgia | **36 (60.0%)** |
|  |  | 11b.Joint Pain | **35 (58.3%)** |
| 12. | Dermatological | 12a.Severe Rash | 5 (8.3%) |
|  |  | 12b.Frequent Rash | 9 (15.0%) |
| 13. | Gastrointestinal | 13a.Nausea | 15 (25.0%) |

All symptoms greater than 20% is show in bold

Suppl Table 2: Frequency of SARS CoV2 positivity in fecal samples in a longitudinal follow-up

| TYPE OF SHEDDERS | n = 60 | Baseline | 6 Month | 12 Month |
| --- | --- | --- | --- | --- |
| Prolonged Shedders | 23 | 23 | 00 | 00 |
| Persistent shedders | 07 | 07 | 07 | 02 |
| Intermittent shedders | 04 | 00 | 02 | 02 |
| Non shedders | 26 | 00 | 00 | 00 |
| Total |  | **30** | **09** | **04** |
